# Sleep and Alcohol Use Patterns During Federal Holidays in the United States

**DOI:** 10.1101/2021.11.04.21264353

**Authors:** Rachel M Heacock, Emily R Capodilupo, Mark É Czeisler, Matthew D Weaver, Charles A Czeisler, Mark E Howard, Shantha MW Rajaratnam

## Abstract

We conducted a retrospective observational study using remote wearable and mobile application data to identify US public holidays associated with significant changes in sleep behaviors, including sleep duration, bedtime and waketime, and the consistency of sleep timing, as well as changes in the point prevalence of alcohol use. These metrics were collected and analyzed from objective, high resolution sleep-wake data and survey responses of 24,250 US subscribers to the wrist-worn biometric device platform, WHOOP (Boston, Massachusetts, USA), who were active users during May 1, 2020 through May 1, 2021. Compared to baseline, statistically significant differences in sleep and alcohol measures were found on the US public holidays and their eves. For example, New Year’s Eve corresponded with a sleep consistency decrease of 13.8% (± 0.3), a sleep onset of 88.9 minutes (± 3.2) later, a sleep offset of 78.1 minutes (± 3.1), and more than twice as many participants reported alcohol consumption (138.0% ± 6.7) compared to baseline. The majority of US public holidays and holiday eves were associated with sample-level increases in sleep duration, decreases in sleep consistency, later sleep onset and offset, and increases in the prevalence of alcohol consumption.

**Significance Statement:** US public holidays were associated with increased sleep duration, decreased sleep consistency, and later sleep timing among US adult users of an objective, validated commercial fitness tracker. Holidays were also associated with an increased prevalence of participants having reported alcohol use. Given the adverse health impacts of sleep timing variability on both weekends and during the transition to daylight savings time, and of increased alcohol use on weekends and public holidays, further investigation of the health impacts of these behavioral changes is warranted.

## Introduction

In the United States, the government and workplaces observe 10 public holidays annually. In 2020 and 2021, each holiday corresponded to approximately 151 million employed persons receiving a day off from work (1). Changes to sleep associated with time off from work have predominantly been investigated in the context of vacations (2, 3) and not specific holidays, while studies of alcohol use patterns have found that consumption of alcohol among regular drinkers increases on holidays (4) and is substantially elevated on special occasions (5). Daylight savings time transitions are a corollary to holidays. The transition of moving clocks forward by one hour for daylight saving time in the spring contributes to shortened sleep and is associated with increased motor vehicle accidents, workplace injuries, heart attacks, and all-cause mortality (6– 10). However, changes to both sleep and alcohol use on specific holidays over the course of a year have not been comprehensively characterized. As such, we sought to identify US public holidays associated with significant changes in health behaviors, including sleep duration, consistency, and timing, as well as changes in the point prevalence of alcohol use.

Weekly changes in sleep patterns can have a significant impact on overall health. The majority of working individuals change their sleep timing and duration depending on next-day commitments (11); this behavior results in cyclical population-level data patterns in which weekend sleep episodes are longer than weekday sleep episodes. Weekday sleep episodes are frequently of insufficient duration, such that many people accrue sleep debt during the work week, and attempt to compensate by extending the duration of weekend sleep episodes (12). Differences in the timing of sleep on free days and work days can result in a misalignment between an individual’s circadian rhythms and the timing of social constraints (i.e., social jetlag)(13). In these cases, social jetlag results in weekly population-level changes in sleep behavior in which weekend sleep duration is longer and the timing of sleep onset and sleep offset occur later compared to weekday sleep duration and timing. Such increased variability in sleep habits is associated with obesity, two-fold increased risk of metabolic syndrome, and diabetes or prediabetes, among other adverse health consequences (14, 15). Conversely, past research has indicated that longer catch-up sleep episodes on weekends or free-days are sometimes beneficial, particularly among individuals who obtain insufficient sleep on workdays. Compensatory weekend catch-up sleep has been associated with a lower body mass index (BMI)(16)] and lower levels of high-sensitivity C-reactive protein (hsCRP)(17) in such populations. Consistent short sleep duration is associated with an increased risk of mortality, but prospective cohort studies have found that those who compensate for shorter sleep episodes during the week with longer sleep duration over the weekend do not have an increased risk of mortality (18). Like weekends, holiday eves that fall on weekdays can provide a longer window of sleep opportunity and greater ability to self-select sleep timing in the absence of work.

As with sleep, weekly differences in alcohol use patterns have been reported, with more consumption on weekends compared to weekdays (4, 19). Such increases have been temporally associated with increased occurrence of motor vehicle accidents among young Swiss men, with parallel peaks in alcohol consumption and alcohol-related motor vehicle accidents on Fridays and Saturdays, as well as an increase on public holidays and the night before (20). Similarly, motor vehicle accidents in the US peak on Independence Day (July 4), and a high portion of alcohol-related crashes and elevated alcohol impairment-related pedestrian crash deaths occur on New Year’s Day (January 1)(21). Moreover, the combination of legal low-dose alcohol and altered sleep (prolonged wake or sleep restriction) are synergistic, resulting in worse impairment than observed at higher alcohol levels that have been shown to increase accident risk (22, 23). More broadly, alcohol use, particularly heavy alcohol consumption, is a risk factor for myriad adverse health consequences, including infectious diseases, diabetes, mental health and substance use conditions, liver and pancreatic diseases, and serious injuries (24). Understanding population-level changes in alcohol use patterns may therefore inform public health efforts to reduce the occurrence of driving under the influence or operating while impaired.

This analysis aimed to investigate the magnitude of changes to sleep duration, consistency, timing, and the point prevalence of alcohol consumption on and prior to public holidays in the US. The findings demonstrate which holidays are associated with the greatest changes from typical sleep and alcohol use patterns among adults, which may have public health implications.

## Results

In total, 24,250 participants met the inclusion criteria and were included in the primary analytic sample (with objective sleep-wake data for an average of 321 out of 365 [87.9%] days), and 13,904 participants were included in the alcohol subsample.

Across participants, 10,350,760 sleep episodes and 5,777,008 alcohol survey responses were recorded and analyzed. Participants (n=24,250) were on average aged 37.6 (± 9.8) years. Males (n=18,060, 74.5% of sample) had an average age of 37.9 (± 9.8) years and females (n=6,187, 25.5% of sample) had an average age of 37.0 (± 9.6) years. Participants who reported sufficient information on alcohol (n=13,904) had an average age of 37.8 (± 9.7) years. Additional demographic information such as race and socioeconomic status were not available.

Holidays and their eves represented several local minima and maxima in sleep and alcohol data (with other days associated with marked changes in sleep and alcohol use including Daylight Savings Time [November 1, 2020] and the US Presidential Election [November 3, 2020])(Figure 1). The holidays and their eves also stand apart from surrounding weekends, indicating an effect separate from that of a standard day off from work. Nearly all holidays were significantly different from baseline in each metric.

**Figure 1.**
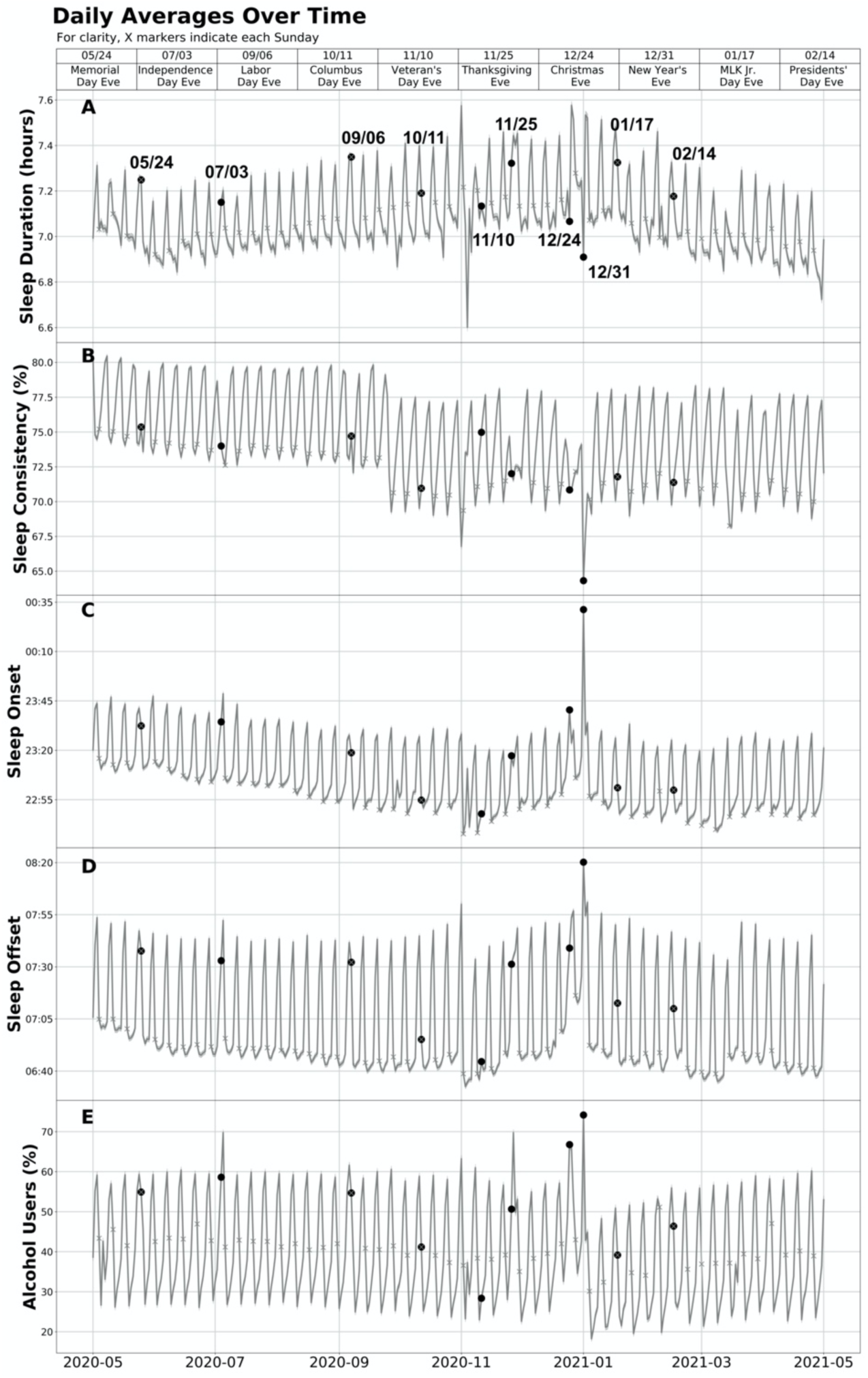
Daily Averages Over Time. Data were collected on participants’ sleep duration, sleep consistency, sleep timing, and alcohol use from May 1, 2020 through May 1, 2021. This figure shows the daily sample means and 95% confidence intervals over the study interval for sleep duration, consistency, and timing. The measure for alcohol is the percentage of users who responded affirmatively to having used alcohol in daily surveys. Each round scatter point represents the eve of a US public holiday and indicates a nocturnal sleep episode that ended the following day. For example, the point on 12/31/20 (New Year’s Eve) represents the nocturnal sleep episode that took place from the evening of December 31, 2020 (or the early morning hours of January 1, 2021) through the morning of January 1, 2021.

### Sleep Duration (Figure 2A)

The mean absolute change from baseline sleep duration during US public holidays during the study interval was 1.9%. Including holiday eves, the absolute change was 2.1%. 15 of the 20 (75%) holidays and eves were associated with significant increases in average sleep duration, with an average increase of 2.5%. There were significant decreases in sleep duration on two of the 20 (10%) holidays and eves, with an average decrease of 1.4%. The largest average increases from baseline sleep duration occurred on Thanksgiving Day (5.2% ± 0.3), Christmas Day (5.1% ± 0.3), and New Year’s Day (4.8% ± 0.3). The largest average decrease from baseline sleep duration occurred on New Year’s Eve (−2.5% ± 0.3), with Memorial Day (−0.3% ± 0.2) as the only other public holiday with a significant decrease in sleep duration. Changes to sleep duration on Christmas Eve, Columbus Day, and Veterans Day were not significant (P > 0.05).

**Figure 2.**
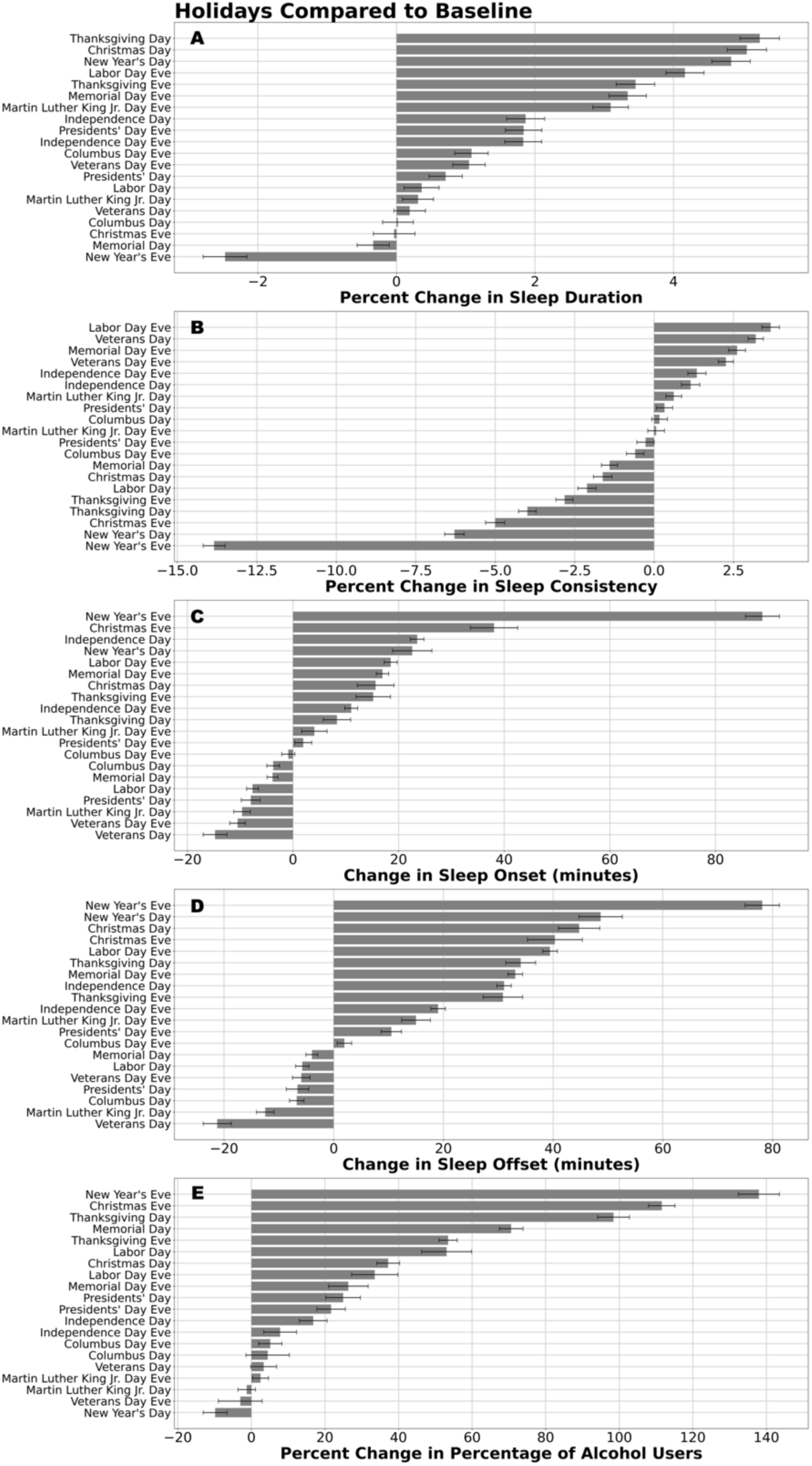
Holidays Compared to Baseline. The percent change from baseline sleep duration and sleep consistency, change in minutes of sleep onset and offset timing, and percent change in the percentage of participants reporting alcohol use were calculated across the sample for each of the US public holidays and eves are shown. For each measure, holidays and eves were ranked from the largest increase to the largest decrease compared to baseline sleep and alcohol use measures. Error bars on sleep duration, consistency, and timing panels represent 95% confidence intervals. Error bars on the alcohol panel represent the propagation of error from binomial proportion confidence intervals of the holidays and their baselines.

### Sleep Consistency (Figure 2B)

The mean absolute change from baseline sleep consistency during US public holidays was 2.1%. Including eves, the absolute change was 2.8%. The eight holidays with significant increases in sleep consistency had an average increase of 1.9%. Nine of 20 (45%) holidays and eves were associated with significant decreases in sleep consistency, with an average decrease of 4.2%. New Year’s Eve had the largest change (−13.8% ± 0.3). The next largest change was on Christmas Eve (−6.3%). The night before Labor Day had the highest average increase in sleep consistency from baseline (2.6%). Sleep consistency differences spanned a larger range (20.2%) when compared to sleep duration differences (16.5%). The percent changes in sleep consistency on Columbus Day and the night before Martin Luther King Jr Day were not significant (P > 0.05).

### Sleep Onset (Figure 2C)

The mean absolute change from baseline in the timing of sleep onset was 11.8 minutes on public holidays. Including their eves, sleep onset changed on average 16.2 minutes. The majority of changes in sleep onset timing (12 of 20 [60%]) were delays in sleep onset, indicating a population-level bedtime that was, on average, 22.1 minutes later on those holidays. The largest average delay in sleep onset was on New Year’s Eve, on which the average change was an 88.9-minute (± 3.2) delay in sleep onset time. Seven of the 20 (35%) holidays and eves were associated with significantly earlier sleep onset times compared to baseline – on average 8.3 minutes earlier. Each of the holidays that had an earlier average time of sleep onset, indicating an earlier average bedtime, were on Monday holidays.

### Sleep Offset (Figure 2D)

The mean absolute change from baseline in the timing of sleep offset on public holidays was 21.5 minutes. This was almost twice the average change of sleep onset timing. Including the holiday eves, the average change was 24.5 minutes. 13 of 20 (65%) holidays and eves were associated with a later sleep offset–on average 32.8 minutes later. On New Year’s Eve (the morning of New Year’s Day), offset timing delayed 78.1 minutes (± 3.1) later, the largest change in sleep offset. We observed earlier sleep offsets (on average 8.9 minutes earlier) on seven of the 20 (35%) holidays and eves, each of which were Monday holidays.

### Alcohol (Figure 2E)

The mean absolute change from the baseline alcohol use on holidays was 32.0%. Including their eves, the mean absolute change was 36.8%. We observed a significantly increased prevalence of alcohol consumption on 14 of 20 (70%) holidays and eves, with an average increase of 46.7%. The majority of significant changes in the number of alcohol consumers were increases, with one exception of New Year’s Day, when alcohol consumption averaged 9.9% lower than surrounding same days of the week. More than twice as many users reported alcohol consumption on New Year’s Eve (138.0% ± 6.7) and Christmas Eve (111.6% ± 4.7) compared to the baseline.

## Discussion

We found that sleep and alcohol use were significantly different from baseline on the majority of US public holidays and their preceding days among a large (n=24250) US-based sample of adults aged 18 to 65 years who were regular users of the wearable biometric device platform WHOOP. These objective data from year-long recordings using a regularly worn validated device reveal for the first time that US public holidays are associated with significant changes in sleep duration, consistency, and timing compared to baseline sleep patterns. Overall, we found that most holidays and their eves were associated with increased sleep duration, decreased sleep consistency, later bedtimes and wake times (with the exception of Monday holidays), and increased prevalence of alcohol use. The largest average increase from baseline sleep duration occurred on Thanksgiving Day (5.2% ± 0.3). New Year’s Eve was associated with the largest change in sleep consistency, a decrease of 13.8% (± 0.3), and the largest change in sleep onset, which occurred an average of 88.9 (± 3.2) minutes later. Participants also had an average sleep offset 78.1 (± 3.1) minutes later on New Year’s Eve (the morning of New Year’s Day). More than twice as many participants reported alcohol consumption on New Year’s Eve (138.0% ± 6.7) and Christmas Eve (111.6% ± 4.7) when compared to baseline.

The largest reported increase in alcohol consumption prevalence and the largest average decrease in both sleep duration (Figure 2A) and sleep consistency (Figure 2B) occurred on New Year’s Eve, when sleep timing was shifted later by more than one hour. The second largest delay in sleep offset occurred on New Year’s Day. Notably, the largest decrease in alcohol consumption prevalence relative to baseline also occurred on New Year’s Day, which followed the largest increase in prevalence that was observed on the previous day. Thanksgiving Day also stood out from the rest of the holidays with the largest average increase in sleep duration, with the third highest increase in alcohol consumption prevalence and fourth largest decrease in sleep consistency. Interestingly, the majority of the holidays that were associated with a decrease in sleep consistency were winter holidays. Lastly, many of the holidays and eves are associated with significant increases in sleep duration that is larger than the effect of melatonin on patients with sleep disorders (+8.25 minutes [95% CI 1.74 to 14.75], p = 0.013)(25) and the impact of Cognitive Behavioral Therapy for Insomnia (CBT-I) (+7.61 minutes, [95% CI -0.51 to 15.74])(26).

## Strengths and Limitations

Strengths of this study include the large sample size and the collection of objective, high-resolution (i.e., continuous 1 Hz recordings to generate 30-second epoch resolution sleep stage estimates), and largely complete (i.e., minimal missingness, with data for an average of 87.9% of the study interval) sleep-wake data from a sleep wearable device validated against polysomnography (27). Limitations of this study may include self-reported alcohol data, which could be subject to social desirability and other biases. For alcohol data in particular, as survey responses were opt-in for subscribers, subscribers who rarely drink alcohol may have been less likely to track their alcohol use. The data shown should be interpreted as relative drinking prevalence among those who drink, which may overrepresent the true prevalence of population-level drinking. On the other hand, if the findings in our sample were generalizable, the increased prevalence on holidays may underestimate the population-level increase. Demographic characteristics outside of gender and age were not collected, so the extent to which the sample is representative of the US adult population is unknown. Given that WHOOP is a personalized digital fitness and health platform, participants may be more informed about sleep and its impact on performance compared to the general population. Finally, the study interval analyzed occurred entirely during the coronavirus disease 2019 (COVID-19) pandemic, and the associated stay-at-home orders and social distancing mandates may have impacted the behavioral changes ordinarily associated with holiday observance; however, population movement increased following the lifting of the first stay-at-home order, which occurred prior to the start of the study interval (April 24, 2020), suggesting that the peak impact of mobility restrictions due to the pandemic had passed (28). Further research following the complete lifting of social distancing mandates will be required to understand how these data were impacted by the present mandates.

Overall, we found that most US public holidays were associated with significant changes in sleep duration, sleep consistency, and sleep timing among a large sample of wearers of a commercial fitness tracker. Holidays were also associated with changes in point prevalence of alcohol use. Similar changes to sleep and alcohol consumption have been associated with adverse health impacts. As such, this research warrants further investigation of the behavioral changes associated with public holidays.

## Materials and Methods

Data were analyzed retrospectively from subscribers to the wearable biometric device platform, WHOOP, Inc. Sleep onset, offset, duration, and architecture were captured using a wrist-worn multi-sensor (tri-axial accelerometer, optical heart rate sensor, capacitive touch sensor and ambient temperature sensor) device (WHOOP Inc, Boston, Massachusetts, USA).25 WHOOP has been validated against polysomnography (27, 29, 30) and has demonstrated acceptable two-stage (wake or sleep) categorization to automatically detect sleep, with 86% agreement with polysomnography (29, 30). Subscribers were also prompted in optional daily surveys delivered through the WHOOP mobile application, which included a question on alcohol intake during the previous day, reported as a binary (yes/no). Holidays analyzed included all 10 US public holidays as observed by the Federal Reserve System (31): Memorial Day (May 25, 2020), Independence Day (also known as Fourth of July; July 4, 2020), Labor Day (September 7, 2020), Columbus Day (also known as Indigenous Peoples’ Day or Indigenous Peoples Day; October 12, 2020), Veterans Day (November 11, 2020), Thanksgiving Day (November 26, 2020), Christmas Day (December 25, 2020), New Year’s Day (January 1, 2021), Martin Luther King Jr Day (also known as Martin Luther King Junior’s Birthday; January 18, 2021), and Presidents’ Day (also known as Washington’s Birthday; February 15, 2021).

The inclusion criteria for the primary analytic sample (sleep data) were: (1) being a US-based member of the WHOOP platform who collected data for 255 (70%) or more days out of the 365-day study interval from May 1, 2020 through May 1, 2021; and (2) being between the ages of 18 and 65 years for the entirety of the data collection interval. From the primary analytic sample, a subsample of participants who responded to optional daily surveys about alcohol consumption for 255 (70%) or more days and all holidays during the data collection interval were included in the alcohol use subsample.

The age range chosen represents working-age individuals, who are most likely to have their work schedules impacted by public holidays (32). Participants were chosen from only the US to avoid confounding the data with international differences in holiday observances. Participants without sleep-wake data on any of the public holidays were excluded from the analysis. Additionally, individual sleep episodes for which complete data were not available (for example, due to loss of device charge) were not counted towards participants’ required minimum of 70% compliance for inclusion.

The Monash University Human Research Ethics Committee reviewed and approved the study protocol. All participants in the study consented at registration with WHOOP to the use of their data for the purposes of scientific research and could withdraw consent at any time.

## Statistical Analysis

Sleep duration, sleep consistency (a proprietary metric of the WHOOP platform adapted from the Sleep Regularity Index (33) to account for recency in the weights of comparator sleep episodes) (34), sleep timing, and alcohol data were analyzed individually. To determine a baseline for holiday comparisons, sleep and alcohol data from the same day of week for four weeks preceding and four weeks following the date of each public holiday were averaged. For example, data for Thanksgiving Day (Thursday, November 26, 2020) were compared to data from the four calendar Thursdays preceding and four following November 26. The baselines were comprised of same day of week to account for population-level variance in sleep behaviors based on day of week (12). A local rather than a complete-year baseline was selected to avoid the confounding effect of seasonal variation in sleep and alcohol use patterns and of the effect of the fluctuating severity of the COVID-19 pandemic and its associated physical distancing mandates (12, 34).

Days preceding each holiday (herein, eves) were analyzed separately to account for the potential impacts on both sleep and behavior leading into as well as occurring on a holiday. For example, we hypothesized that in anticipation of Labor Day, which occurs on a Monday, the preceding Sunday night might have higher prevalence of alcohol consumption, shifted sleep timing, and overall extended sleep relative to a typical Sunday night, but Labor Day itself may resemble a non-holiday Monday night in which sleep and alcohol patterns are indifferent from typical pre-work night patterns. Christmas Eve and Day and New Year’s Eve and Day are exactly one week apart and therefore a necessary exception to the described baseline methodology. These days were excluded from each other’s baseline calculations in order to confounding the baseline with another holiday.

Daily sleep duration, consistency (scored on a 0 to 100 scale), and timing were calculated for participants over the year-long study interval. An average by day was taken across participants for each sleep metric to visualize the variance in metrics over the duration of the study. Sleep episodes concluding in the morning of one day were assigned to the previous day’s sleep onset date. For example, sleep episodes concluding the morning of January 2, 2020 were treated as if the sleep episode were initiated on date January 1, 2020. If the sleep episode initiated in the early morning hours of January 2, 2020, the episode was still assigned to January 1, 2020. The average values for each participant were compared to their individual baseline values and used to calculate percent changes for sleep duration and sleep consistency for both the public holidays and their eves. Population estimates for percent change were created by averaging the participant estimates. The 95% confidence intervals based on between-subjects variability were estimated as well.

The alcohol data were analyzed using the daily percentage of subscribers who reported alcohol consumption in the optional survey out of the total subscribers who responded to the question. Individuals included in this subset of analysis who did not respond on a given day were omitted from the sample for that day. Similar to the method of analyzing sleep data, a baseline using the same day of week from four weeks prior to and four weeks after each holiday was created by taking an average of the eight days. The binomial proportion confidence interval was calculated for both the holidays and their baselines. The percent change from baseline was calculated for both the public holidays and their eves. The propagation of error for the percent change was computed, accounting for the correlation of measurement error and correlation of uncertainties in the group averages. The changes in individual measures (sleep duration, sleep consistency, sleep onset, sleep offset, and alcohol use) for each holiday and eve were then ranked among the holidays and eves in each measure from most positive to most negative change compared to baseline.

All analyses were conducted using the Python programming language version 3.6.2 (Python Software Foundation). The threshold for statistical significance was set at 0.05.

## Data Availability

Public data sharing for these purposes is ethically and legally restricted. The data is collected and used with the consent of individuals who purchase a WHOOP membership (collectively, "WHOOP Members") and agree to the WHOOP terms of use (https://www.whoop.com/termsofuse/) and privacy policy (https://www.whoop.com/privacy/full-privacy-policy/). While WHOOP terms of use and privacy policy permit WHOOP to use collected data that has been aggregated or de-identified in a manner that no longer identifies an individual for informational purposes, analytics or WHOOP's own research purposes, the consent obtained from WHOOP Members does not extend to making the data publicly available for a third party to use for its own purposes. As such, WHOOP's legal department will not permit the data to be shared for these purposes.

## Acknowledgments

M.É.C. gratefully acknowledges funding by The Kinghorn Foundation through a 2020 to 2021 Australian-American Fulbright Scholarship.

